# Congenital Heart Disease: Assessing Viral Etiology in infants attending at a tertiary care hospital

**DOI:** 10.1101/2023.04.19.23288543

**Authors:** Syeda Fakiha Mehreen, Mote Srinath, Waseema Sultana, Vannavada Sudha Rani

## Abstract

**Background:** Congenital heart disease (CHD) is the commonest birth defect worldwide, affecting millions of newborns every year. Limited information is available regarding risk factors for its causation.

**Objectives:** This study was planned with an aim to determine the viral etiology in infants with congenital heart disease. Although a number of viruses diseases have been etiologically linked to congenital defects, only two viruses namely rubella virus and cytomegalovirus are definitely proved to be associated with anomalies in infants.

**Materials and Methods:** This Prospective study was conducted at a paediatric tertiary care hospital for a period of 6 months. Hundred (100) Infants with structural Congenital heart defects (CHD) based on Echocardiography findings were included in the study after taking informed consent from their mothers. Serum samples were collected and sent to Virology lab. The samples were tested for Rubella and Cytomegalovirus IgM antibodies using Enzyme Linked Immunosorbent Assay (ELISA).

**Results:** Out of 100 infants with congenital heart defects, (M: F:56:44) 4% were seropositive for rubella IgM antibodies whereas 20% showed seropositivity of Cytomegalovirus IgM antibodies. Median age of infants was 25 days. The most common congenital heart defect in Rubella positive infants was Patent ductus arteriosus followed by Atrial septal defect whereas it was Patent ductus arteriosus only in CMV positive infants followed by Patent Foramen ovale. 6% of infants died and among them 50% had infection with CMV while 11% showed infection with Rubella.

**Conclusion:** Regular screening by TORCH (Toxoplasmosis, Rubella, Cytomegalovirus, HSV) panel testing of mothers in first trimester is necessary for early detection. Early vaccination strategies should be implemented. Rapid and correct diagnosis of congenital CMV and Rubella infections in infants is very important for the correct therapy selection and proper management of the cases.

## INTRODUCTION

Congenital heart disease (CHD) is the commonest birth defect worldwide, affecting millions of new-borns every year. CHD is typically defined as a structural abnormality of the heart and/or great vessels that is present at birth.^1^ Considering a birth prevalence as 9/1000, the estimated number of children born with CHD every year in India approximates 240,000, posing a tremendous challenge for the families, society and health care system.^2^ Most congenital heart lesions are probably not the result of genetic or environmental factors acting alone, but rather the result of these factors acting in concert with one another. The genetically predisposed foetus is exposed to the appropriate environmental factor at a particular stage during organogenesis which leads to the development of malformations.^3^

Even after advancement in medicine today, exact aetiology of CHD remains unexplained or multifactorial. Very little information is available regarding causative modifiable risk factors. Some viruses, such as rubella and human cytomegalovirus, are known to cross the placental barrier and infect the foetus. In other cases of maternal viral infections, such as herpes simplex, evidence for transplacental passage is less convincing and foetal damage or neonatal disease may be coincidental or associated with perinatal infection. Although several maternal viral diseases have been etiologically incriminated in congenital defects, only two-rubella and cytomegalovirus infection-are definitely proved to be associated with anomalies or mental retardation in infants.^4^

Congenital rubella syndrome (CRS) has long been associated with the triad of deafness, cataract, and cardiovascular malformations (CVMs) (Gregg, 1945). CRS continues to be a common cause of CHD in the developing countries as rubella vaccination is not a part of routine immunisation yet. However, as the diagnosis of congenital CVMs has been based often on clinical examination alone, the exact nature of the spectrum of cardiac phenotypes associated with CRS has not been well established.^5^ Even after implementing measles-rubella (MR) vaccination in routine childhood national immunization programs, a significant proportion of young adult women remain susceptible to rubella infection for a period, responsible for continuing CRS cases.^6^

## MATERIALS AND METHODS

In this study, we included 100 infants with congenital abnormalities, attending a paediatric tertiary care hospital. The serum samples were collected and stored at -20 until processed. Demographic details and clinical features were recorded. 2D Echocardiography and clinical data was utilised to establish diagnosis of congenital heart disease.

The inclusion criteria for the study included infants with Congenital birth defects like congenital heart disease, bilateral cataract, microcephaly, conjugated hyperbilirubinemia, hydrocephaly and anotia. Infants without any congenital abnormalities were excluded. The samples were screened for Rubella and Cytomegalovirus IgM antibodies using Enzyme Linked Immunosorbent Assay (ELISA) (Diapro Diagnostics ELISA kit). The assay is based on the principle of “IgM capture” where IgM class antibodies in the sample are first captured by the solid phase coated with anti hIgM antibody. Results were interpreted based on controls that were provided with the kit. An Infant sample was said to be positive for IgM when its absorbance value was higher than the absorbance value of the cut-off control. Positivity of IgM antibodies against CMV and Rubella in a sample indicates active infection of CMV and Rubella respectively. The study was completed with all the data analyzed and tabulated. The study was approved by the Institutional ethics Committee, Osmania medical College, Hyderabad, Telangana.

## RESULTS

A total of 100 infants were tested for Rubella and Cytomegalovirus IgM antibodies. This included 56 male infants and 44 females. 88% of infants belonged to rural area when compared to 12% of infants in urban areas with congenital birth defects. The age range of infants was 0-11 months, median age being 25 days.

The percentage of patients with IgM seropositivity for rubella is 4%, three females and one male were positive for Rubella IgM antibodies. Median birthweight was 2.35 kg. All the patients showed congenital heart disease with Patent ductus arteriosus associated with Atrial septal Defect, one male infant had conjugated hyperbilirubinemia, ventricular septal defect along with microcephaly. Three female patients had congenital heart disease with atrial septal defect, ventricular septal defect, and patent ductus arteriosus as shown in Table 1.

**Table 1.** Seropositivity of Rubella with Congenital Heart Disease.

| S.no | Rubella positive (n=4) | Age (Days) | Birthweight (kg) | Sex | CONGENITAL ABNORMALITIES |  |  |  |  |  |  |  |
| --- | --- | --- | --- | --- | --- | --- | --- | --- | --- | --- | --- | --- |
|  |  |  |  |  | ASD | VSD | PDA | PFO | HL | BCC | CHB | Microcephaly |
| 1 | PATIENT 1 | 1-20 | 2.5-3.0 | F | ] | ] | ] |  |  |  | ] | ] |
| 2 | PATIENT 2 | 21-40 | 2.0-2.5 | F | ] |  | ] | ] |  |  |  |  |
| 3 | PATIENT 3 | 1-20 | 2.0-2.5 | F | ] | ] | ] |  |  |  |  |  |
| 4 | PATIENT 4 | 21-40 | 2.5-3.0 | M | ] | ] |  |  |  |  | ] | ] |

\*ASD – ATRIAL SEPTAL DEFECT, VSD – VENTRICULAR SEPTAL DEFECT, PDA – PATENT DUCTUS ARTERIOSUS, PFO – PATENT FORAMEN OVALE, HL- HEARING LOSS, BCC – BILATERAL CONGENITAL CATARACT, CHB – CONJUGATED HYPER BILIRUBINEMIA

The percentage of patients with IgM seropositivity for CMV is 20%. Out of 20, 9 females and 11 males were positive for Cytomegalovirus antibodies. Median birthweight was 2.5 kg. Most of the patients positive for CMV had CHD with Patent Ductus Arteriosus. Two males and one female had articular septal defect. Three females and one male had ventricular septal defect. Four females and two males had Patent foramen ovale. One male infant represented moderate hearing loss, one male and one female infant had bilateral congenital cataract and Microcornea respectively as shown in Table 2.

**Table 2.**
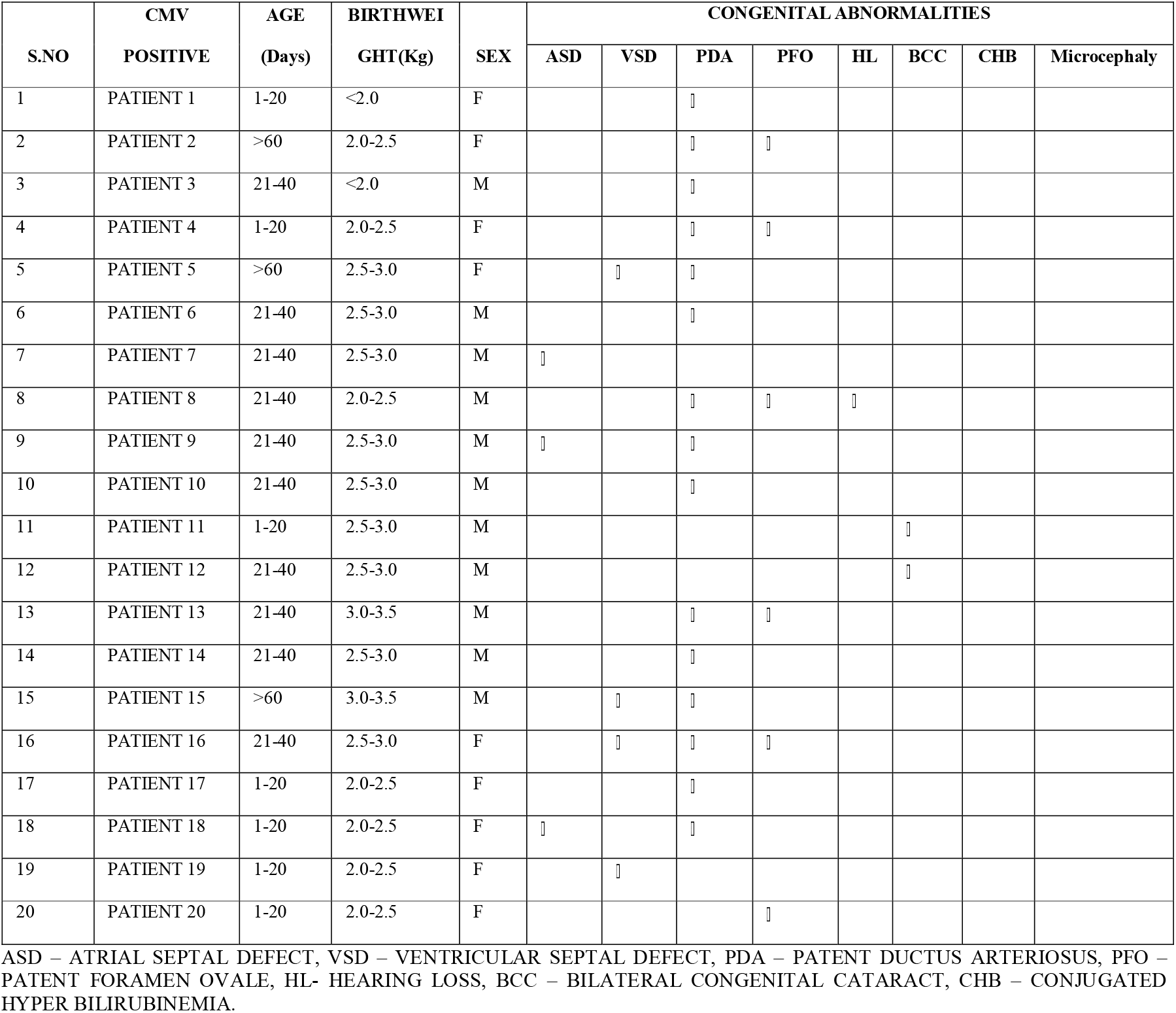
Seropositivity of CMV with Congenital Heart Disease:

| S.NO | CMV<br>POSITIVE | AGE<br>(Days) | BIRTHWEI<br>GHT(Kg) | SEX | CONGENITAL ABNORMALITIES |  |  |  |  |  |  |  |
| --- | --- | --- | --- | --- | --- | --- | --- | --- | --- | --- | --- | --- |
|  |  |  |  |  | ASD | VSD | PDA | PFO | HL | BCC | CHB | Microcephaly |
| 1 | PATIENT 1 | 1-20 | <2.0 | F |  |  | ] |  |  |  |  |  |
| 2 | PATIENT 2 | >60 | 2.0-2.5 | F |  |  | ] | ] |  |  |  |  |
| 3 | PATIENT 3 | 21-40 | <2.0 | M |  |  | ] |  |  |  |  |  |
| 4 | PATIENT 4 | 1-20 | 2.0-2.5 | F |  |  | ] | ] |  |  |  |  |
| 5 | PATIENT 5 | >60 | 2.5-3.0 | F |  | ] | ] |  |  |  |  |  |
| 6 | PATIENT 6 | 21-40 | 2.5-3.0 | M |  |  | ] |  |  |  |  |  |
| 7 | PATIENT 7 | 21-40 | 2.5-3.0 | M | ] |  |  |  |  |  |  |  |
| 8 | PATIENT 8 | 21-40 | 2.0-2.5 | M |  |  | ] | ] | ] |  |  |  |
| 9 | PATIENT 9 | 21-40 | 2.5-3.0 | M | ] |  | ] |  |  |  |  |  |
| 10 | PATIENT 10 | 21-40 | 2.5-3.0 | M |  |  | ] |  |  |  |  |  |
| 11 | PATIENT 11 | 1-20 | 2.5-3.0 | M |  |  |  |  |  | ] |  |  |
| 12 | PATIENT 12 | 21-40 | 2.5-3.0 | M |  |  |  |  |  | ] |  |  |
| 13 | PATIENT 13 | 21-40 | 3.0-3.5 | M |  |  | ] | ] |  |  |  |  |
| 14 | PATIENT 14 | 21-40 | 2.5-3.0 | M |  |  | ] |  |  |  |  |  |
| 15 | PATIENT 15 | >60 | 3.0-3.5 | M |  | ] | ] |  |  |  |  |  |
| 16 | PATIENT 16 | 21-40 | 2.5-3.0 | F |  | ] | ] | ] |  |  |  |  |
| 17 | PATIENT 17 | 1-20 | 2.0-2.5 | F |  |  | ] |  |  |  |  |  |
| 18 | PATIENT 18 | 1-20 | 2.0-2.5 | F | ] |  | ] |  |  |  |  |  |
| 19 | PATIENT 19 | 1-20 | 2.0-2.5 | F |  | ] |  |  |  |  |  |  |
| 20 | PATIENT 20 | 1-20 | 2.0-2.5 | F |  |  |  | ] |  |  |  |  |

ASD – ATRIAL SEPTAL DEFECT, VSD – VENTRICULAR SEPTAL DEFECT, PDA – PATENT DUCTUS ARTERIOSUS, PFO – PATENT FORAMEN OVALE, HL- HEARING LOSS, BCC – BILATERAL CONGENITAL CATARACT, CHB – CONJUGATED HYPER BILIRUBINEMIA.

Among the 100, 6 infants died, out of which 3 showed presence of CMV IgM antibodies and 1 showed Rubella IgM antibodies. Five (5) of them were diagnosed with patent ductus arteriosus with left to right shunt while one was diagnosed with a large VSD.

## DISCUSSION

When deciding how to classify and analyse heart defects in etiologic studies, investigators need to balance epidemiologic, anatomic, and developmental considerations, with the goal of producing unbiased and precise risk estimates, optimizing statistical power, and, ultimately, providing information useful for intervention and prevention.^7^ Rubella had always been a significant contributor to the causation of CHD.^8^ Rubella is the only viral agent which has been proved to be a true teratogen, which results in congenital malformations.^3^ It is a noncytolytic virus in certain tissues; that is, it does not destroy the cells in which it replicates. In our study, 4% of the infants were positive for rubella virus which was similar to a study by Sanjay Gupta et al. (8.75%). The most common cardiovascular malformation associated with rubella positive infants in our study was PDA (75%) followed by ASD then VSD, similar to a study by Toizumi *et al* who studied 38 CRS infants, among whom PDA (72%) was the most common defect.^9^ This was in contrast to a study by Oster et al where Branch pulmonary stenosis was more common followed by PDA.^5^ Direct viral damage to the septa of the heart may be the cause of the increased incidence of septal defects.^9,10^

Congenital Rubella syndrome can result in prematurity and low birth weight ^11^, and prematurity in turn often results in PDA. In our study median birthweight of infants with Rubella seropositivity was 2.35kg which is low birth weight. ^12,13^ After rubella the best attested example of a virus which affects the unborn baby is cytomegalovirus. For most other viruses the information is at best scanty and at worst merely anecdotal.^14^ Human cytomegalovirus appears to produce no demonstrable illness in the mother, yet may give rise to severe, frequently fatal disease in the foetus and newborn.^14^ CMV afflicted babies are known to shed virus in various bodily secretions especially urine, blood and throat swab, in some cases, for months and years.^15^ In our Study, out of 100 infants 20% showed seropositivity for CMV IgM antibodies which was similar to a study by Gandhoke et al, where 21.6% of infant showed seropositivity.^15^ In our study, 88% of infants with CHD belonged to rural areas. In rural India, Congenital Heart Diseases are a formidable public health problem. There is lack of prenatal detection of heart defects in rural areas due to limited resources, also parents seek medical attention only when children develop significant symptoms due to lack of information and social stigma.

## CONCLUSION

Congenital Heart defects have become common type of congenital anomalies in new-borns which has emerged as a significant cause of infant mortality. Caring for children with CHD is very expensive and time consuming in developing countries. The Indian government is taking a variety of initiatives to improve children’s health through various programmes and plans that are expected to help children with congenital heart disease, particularly those who are underprivileged and marginalised. It is to be concluded that Rubella and CMV are associated with congenital heart disease. The risk of foetus getting exposed to intrauterine or perinatal infection varies with the type of virus, time of exposure and with the mother’s immune status. TORCH Screening is necessary in the early trimester of pregnancy to rule out maternal infection. Furthermore, Rapid, and correct diagnosis of congenital CMV and Rubella infections in neonates is very important for the correct therapy selection and proper management of the cases. Chemical substances known to have some antiviral activity might be considered useful if begun soon after birth; however, associated side effects might rule them out. In some cases, specific immunoglobulins can be administered as a preventative measure, and antiviral chemotherapy may be necessary. Collaboration between Government authorities, frontline health care workers, paediatricians and other non-profit organisations is necessary to improve the overall outcome for babies with CHD. There is a need for dedicated CHD units with trained medical experts in every district or cluster of the district. Health policies and schemes should be implemented directing more resources and financial support for successful and timely treatment.

## Data Availability

All data produced in the present study are available upon reasonable request to the authors.

## Acknowledgments

The authors acknowledge the funding support from DHR-ICMR and CRS project.

## Conflicts of Interest

None

## Authors contribution

Conceptualization-SFM, MS; Data Collection and testing-WS; Data analysis-SFM, MS; Manuscript Preparation-WS, SFM, MS; Manuscript Review-VSR.

## Notes

### Competing Interest Statement

The authors have declared no competing interest.

### Funding Statement

This study was funded by DHR-ICMR,VRDL for procuring testing kits.

### Author Declarations

The Present study is approved by the Institutional Ethics Committee, Osmania Medical College, Hyderabad, Telangana.

